# Relevance of Seasonal Revaccination against Respiratory Syncytial Virus in Hematology Patients: First Insights in Stem Cell Transplant Recipients

**DOI:** 10.64898/2026.09.08.26362495

**Authors:** Rabah Redjoul, Laurent Softic, Ludovic Cabanne, Gaëlle Sandillon, Christine Robin, Mohamed Ader, Clément Ourghanlian, Karishma Soocheta, Alexandre Soulier, Slim Fourati, Sébastien Maury

## Abstract

Several vaccines against respiratory syncytial virus (RSV) have been commercialized recently. Their immediate immunogenicity in various conditions of immunosuppression, including patients receiving allogeneic hematopoietic stem cell transplantation (HSCT), have been reported in 2025 by different teams, including ours. In immunocompromised patients, the duration of vaccinal humoral protection as well as the relevance of seasonal revaccination is still unknown. By analyzing the kinetics of humoral responses after a single dose of bivalent RSVpreF vaccine over 12 months in 71 HSCT recipients, we show that annual revaccination benefits mostly those primarily vaccinated within the early 18 months after HSCT. We also provide a first evaluation of vaccinal clinical effectiveness in HSCT patients.

---

After HSCT, RSV can lead to severe infection with a particularly poor prognosis.^1^ Three vaccines targeting the RSV prefusion F protein (RSVpreF) have been approved in the United States and Europe for immunocompetent adults aged 60 years or older, based on large randomized trials.^2–4^ In this population, a single vaccine dose is efficacious against RSV-related infections over up to three successive RSV seasons.^5–7^

In immunocompromised patients, including those receiving allogeneic HSCT, we and others recently documented a less efficient immunogenicity of a first vaccine dose, marked by lowered seroconversion rates following immunization by unadjuvanted or adjuvanted vaccines.^8–12^ Importantly, in seroresponders, the duration of protection remains poorly documented, raising the question of whether annual revaccination is necessary. Alternatively, in non-seroresponding patients, the possibility to obtain an improved immunogenicity following revaccination one year later before the next RSV season also warrants evaluation. This last question is of particular interest in HSCT patients, who mount particularly low antibody responses when vaccinated within the early months following transplantation,^9–11^ but also most often experience a progressive reconstitution of favorable immune conditions over time.

To address these concerns, we analyzed the kinetics of the anti-RSVpreF humoral responses over a 12-month follow-up, as well as the effect and potential benefit of revaccination before the next RSV season, in 71 adult recipients of allogeneic HSCT who received a single dose of the bivalent RSVpreF vaccine (Abrysvo™, Pfizer Inc) before the 2024-2025 RSV season. The immunological assay for anti-pre-F RSV IgG quantification was based on the detection of IgG antibodies to RSV preF antigens in serum using a commercially available “Human Anti-RSV-F0 Antibody IgG titer ELISA Assay Kit” (ACRObiosystems) according to the manufacturer’s instructions. The assay is based on a standard indirect ELISA format. For validation and quantitation in IU/ml, serial dilutions of serum, negative and positive controls, and antiserum to Respiratory Syncytial Virus (WHO 1st International Standard, NIBSC 16/284) were used. In a subset of 15 patients included in the present study, we previously reported the strong correlation between RSVpreF antibodies titers and neutralizing antibody titers.^9^

This study was approved by the Paris Hospital Consortium Committee (N° APHP230185) and by national Ethics Committee (N° EUDRACT/ID-RCB: 2023-A01 420-45). Participants provided written informed consent. We followed the guidelines of the Declaration of Helsinki and the Strengthening the Reporting of Observational Studies in Epidemiology (STROBE) reporting guideline. Comparisons of continuous variables were performed using the Mann-Whitney test. Two-sided P values less than 0.05 were considered significant. Analyses were completed in GraphPad Prism 11.

As the interval between HSCT and vaccination has been consistently identified as the major determinant for anti-RSV humoral response in preliminary studies,^11,9,10^ we analyzed post-vaccinal antibody kinetics separately in patients vaccinated early (≤18 months) versus later (>18 months) after HSCT. The characteristics of patients between these two subpopulations are shown in Table 1. As compared to baseline (BL) antibody titers, vaccination resulted in seroconversion (≥4-fold increase at one month) in 14% (6/43) of patients vaccinated early as compared to 63% (17/27) in those vaccinated later after HSCT. The corresponding median fold-increase in antibody titers was 1.0 (IQR 0.7-2.6) versus 9.6 (IQR 1.0-68.9), respectively. Throughout the 1-year follow-up, median antibody fold changes remained stable in each group (Figure 1A).

**Table 1.** Characteristics of patients according to the interval from HSCT to primary RSV vaccination.

|  |  | Patients, No (%) |  |  |
| --- | --- | --- | --- | --- |
|  |  | All (n=71) | Time from HSCT to primary RSV vaccination |  |
|  |  |  | ≤18 mo (n=44) | >18 mo (n=27) |
| Seroconversion at 1 month after primary vaccination<sup>a</sup> | No | 48 (68) | 38 (86) | 10 (37) |
|  | Yes | 23 (32) | 6 (14) | 17 (63) |
| Time from HSCT to primary vaccination, median (IQR), mo |  | 12 (4-31) | 7 (4-10) | 48 (27-53) |
| **Demographics** |  |  |  |  |
| Sex | Male | 48 (68) | 30 (68) | 18 (67) |
|  | Female | 23 (32) | 14 (32) | 9 (33) |
| Age at transplant, median (IQR), y |  | 59 (48-63) | 53 (37-60) | 62 (59-64) |
| **Transplant characteristic** |  |  |  |  |
| Disease | Myeloid malignancy | 44 (62) | 23 (52) | 21 (78) |
|  | Lymphoid malignancy | 17 (24) | 11 (25) | 6 (22) |
|  | Non-malignant | 10 (14) | 10 (23) | 0 (0) |
| Donor type | HLA-identical sibling | 23 (32) | 14 (32) | 9 (33) |
|  | Haplo-identical related | 23 (32) | 16 (36) | 7 (26) |
|  | Unrelated | 25 (36) | 14 (32) | 11 (41) |
| Post-transplant cyclophosphamide | No | 44 (62) | 25 (57) | 19 (70) |
|  | Yes | 27 (38) | 19 (43) | 8 (30) |
| Acute GVHD grading<sup>b</sup> | 0 | 57 (80) | 37 (84) | 20 (74) |
|  | I | 4 (6) | 3 (7) | 1 (4) |
|  | II | 9 (13) | 4 (9) | 5 (18) |
|  | III | 1 (1) | 0 (0) | 1 (4) |
| Chronic GVHD grading<sup>c</sup> | No | 53 (75) | 37 (84) | 16 (59) |
|  | Mild | 3 (4) | 2 (4.5) | 1 (4) |
|  | Moderate | 8 (11) | 2 (4.5) | 6 (22) |
|  | Severe | 7 (10) | 3 (7) | 4 (15) |
| **Immunosuppressants taken in the 3-mo preceding first vaccination** |  |  |  |  |
| Ciclosporin/sirolimus | No | 34 (48) | 11 (25) | 23 (85) |
|  | Yes | 37 (52) | 33 (75) | 4 (15) |
| Systemic steroids | No | 67 (94) | 40 (91) | 27 (100) |
|  | Yes | 4 (6) | 4 (9) | 0 (0) |
| Rituximab | No | 66 (93) | 41 (93) | 25 (93) |
|  | Yes | 5 (7) | 3 (7) | 2 (7) |
| Ruxolitinib | No | 63 (89) | 41 (93) | 22 (81.5) |
|  | Yes | 8 (11) | 3 (7) | 5 (18.5) |
| No. of immunosuppressants taken | 0 | 30 (42) | 11 (25) | 19 (71) |
|  | 1 | 30 (42) | 25 (57) | 5 (18) |
|  | 2 | 10 (15) | 7 (16) | 3 (11) |
|  | 3 | 1 (1) | 1 (2) | 0 (0) |
| **Immune status at first vaccination** |  |  |  |  |
| ≤3-mo IVIg supplementation | No | 54 (76) | 30 (69) | 24 (89) |
|  | Yes | 17 (24) | 14 (32) | 3 (11) |
| Lymphocytes in PB | ≤ 1.4 Giga/L | 40 (56) | 35 (80) | 5 (18.5) |
|  | > 1.4 Giga/L | 31 (44) | 9 (20) | 22 (81.5) |

<sup>a</sup> Defined as 4-fold or greater rise in anti-RSVpreF antibodies in serum.
<sup>b</sup> Grading according to Glucksberg scale.
<sup>c</sup> Grading according to the National Institutes of Health classification global scoring.
**Abbreviations:** HSCT, allogeneic hematopoietic stem cell transplantation; IQR, interquartile range; HLA, human leukocyte antigens; GVHD, graft-versus-host disease; PB, peripheral blood; IVIg, intravenous immunoglobulin.

**Figure 1.**
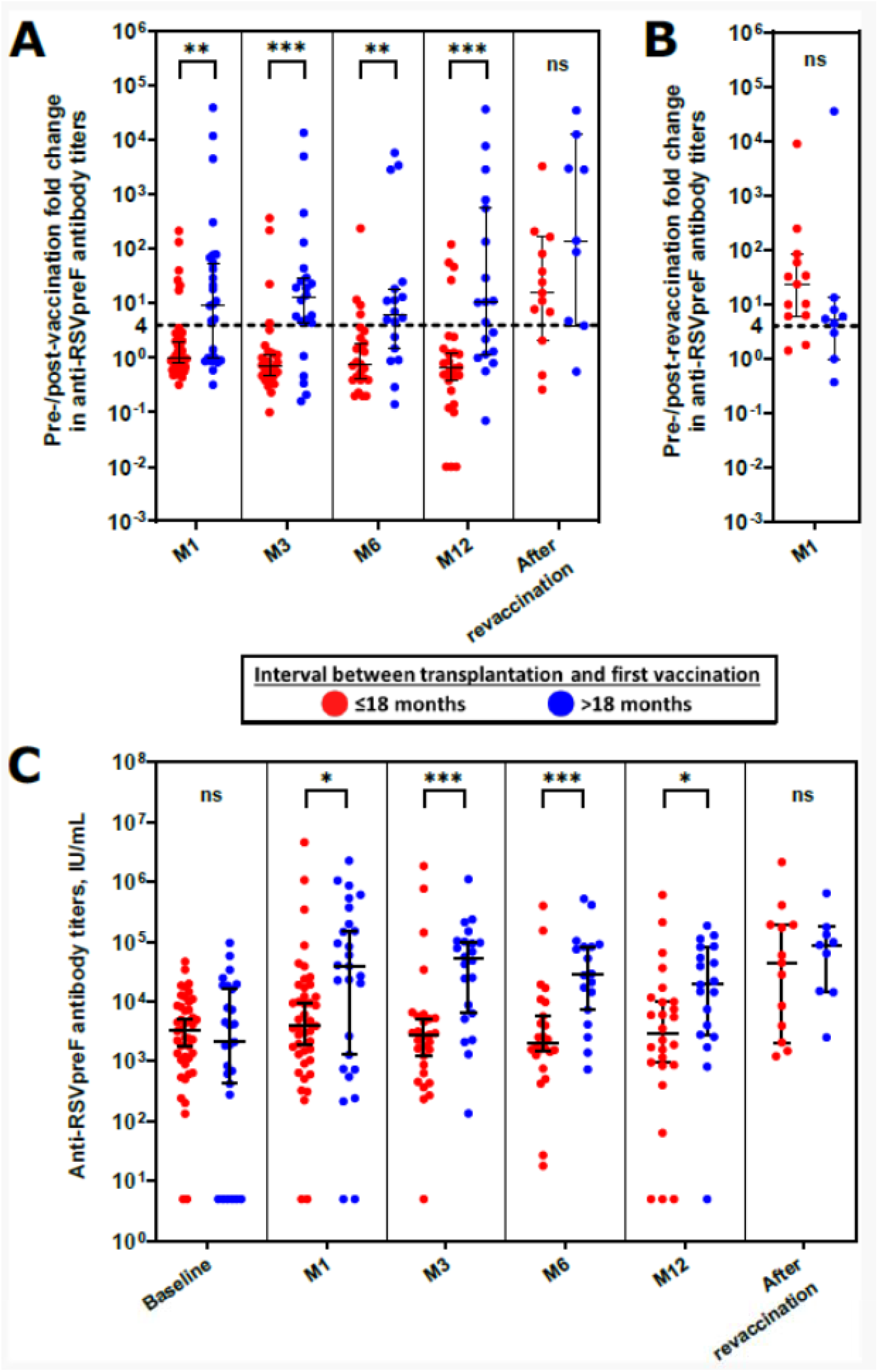
Vaccinal immunogenicity over two RSV seasons. (A) Pre-/post-vaccination fold changes in anti-RSVpreF antibody titers at 1, 3, 6 and 12 months following RSV vaccine (M1 to M12) and after revaccination. At each timepoint, fold changes against baseline pre-vaccination titers are compared between patients primarily vaccinated early (≤18 months, red dots) vs. later after HSCT (>18 months, blue dots). The black dotted line figures the seroconversion threshold of 4-fold increase in anti-RSVpreF antibodies as compared to baseline titers. (B) Pre-/post-revaccination fold changes in anti-RSVpreF antibody titers at one month following RSV revaccination. The black dotted line figures the seroconversion threshold of 4-fold increase. (C) Post-vaccinal kinetics of anti-RSVpreF antibody titers. At each timepoint, titers are compared between patients primarily vaccinated early (≤18 months, red dots) vs. later after HSCT (>18 months, blue dots).
Error bars indicate median 95% CI. Asterisks indicate statistical significance by Mann-Whitney tests (*, *p* < 0.05; **, *p* < 0.01; ***, *p* < 0.001; ns, not significant).

Before the 2025-2026 RSV season, 13 patients of the early-vaccinated and 9 from the late-vaccinated group were revaccinated at a median interval of 12.9 (IQR 11.3-14) and 13.7 (IQR 12.6-14.1) months after the primary 2024-2025 vaccination, respectively. In the former group, revaccination induced a 23-fold increase in anti-RSVpreF antibodies, as compared to 4-fold in the latter (Figure 1B). Compared with baseline levels before the 2024-2025vaccination, both groups showed a ≥4-fold increase in anti-RSVpreF antibodies after revaccination (Figure 1A). Comparable antibody titers were achieved in both groups following revaccination (Figure 1C).

Although the present study was not primarily designed to assess clinical effectiveness, we sought to obtain a preliminary estimate of the impact of RSV vaccination in our department. To do so, we compared the number of PCR-confirmed RSV infections diagnosed in our central lab between i) the pre-vaccination era, i.e., two successive RSV seasons: October 2022 to March 2023 and October 2023 to March 2024) and ii) the post-vaccination era, i.e., two successive RSV seasons: October 2024 to March 2025 and October 2025 to March 2026.

During the pre-vaccination period, 7 out of 58 tested patients (12%) were positive for RSV detection in nasopharyngeal and/or bronchoalveolar samples. During the post-vaccination era, 3 out of 53 tested patients (6%) were positive. Among those 3, two were not vaccinated previously while one was vaccinated one month before documented RSV infection. Two of those 3 previously received rituximab for post-transplant EBV reactivation, including the latter patient who received rituximab 6 months before he was vaccinated against RSV.

Regarding safety, we previously reported adverse events after one single dose of bivalent RSVpreF vaccine in HSCT recipients.^9^ After revaccination in the same cohort, only mild or moderate adverse events were self-reported through structured questionnaires. Neither severe (grade ≥ 3 according to CTCAE v5.0) nor unsolicited adverse events of any grade were reported. Of note, we did not observe any GVHD reactivation or flare neither after RSV vaccination, nor after revaccination one year later.

Limitations of the present study include the small sample size, which precluded multivariable analyses, and the evaluation of only one of the available RSV vaccines. Additional studies are needed to define a clinical protection correlate in these unfavorable immune conditions, also considering the putative impact of cellular responses to vaccination.

Regarding clinical effectiveness of RSV vaccines in transplant recipients, emerging real-world data suggest a lower effectiveness as compared to other situations in immunocompetent patients. A very recent updated meta-analyses in maternal vaccination for infant protection or in adults aged 60 years or older evaluate vaccine effectiveness ≥68% against hospitalization.^13^ By comparison, another recent test-negative case-control study in HSCT recipients also aged 60 years or older reported a clinical effectiveness of 33%.^14^

Our findings suggest that revaccination one year later is particularly beneficial in patients who failed to respond primarily. In these individuals, the response observed after boosting is consistent with progressive reconstitution of adaptive immunity over time after HSCT. Conversely, our findings question the clinical utility of vaccination during the early months after transplantation, when immune recovery is still limited. Within this period of greatest RSV vulnerability, passive immunization with monoclonal antibodies currently available for infant prophylaxis,^16^ may represent a more effective preventive strategy.

In patients primarily vaccinated later after HSCT, the observation of a booster effect at one year indicates that revaccination may also be useful in this subgroup, supporting for a systematic revaccination strategy, while accounting for interindividual variability in immune reconstitution. These results contribute to refining RSV vaccination in immunocompromised persons. Similar studies conducted in other specialized situations such as solid organ transplantation, immune-mediated systemic diseases and/or prolonged immunosuppressive therapies will help tailor patient-adapted vaccination schedules, with the aim to prevent RSV severe infections on the long-term.

## Data Availability

All data produced in the present work are contained in the manuscript

## Acknowledgements

The authors are grateful to Mathieu Leclerc, Vincent Parinet, Cécile Pautas, Selwa Bouledroua and Lydia Roy (all MDs) for patient recruitment; and to Marylene Valmy, Mariline Dos Santos Dominguez, Sophie Lalaque and all the nurses’ staff for their mandatory contribution in blood sample collection. All affiliated to Henri Mondor Hospital, Assistance Publique-Hôpitaux de Paris (AP-HP), Créteil, France.

